# Single-Cell Mendelian Randomization Identifies Cell-Type Specific Genetic Drivers of Lung Cancer Subtypes

**DOI:** 10.64898/2025.11.30.25341293

**Authors:** Qimao Yang, Chengsong Yan, Xiao-fan Wang

## Abstract

**Background:** Genome-wide association studies (GWAS) have identified many loci linked to lung cancer, but connecting these loci to causal genes and relevant cell types remains difficult. Traditional Mendelian randomization (MR) using bulk tissue eQTLs averages signals across diverse cells, masking cell-specific effects. Here, we apply single-cell eQTL based MR to infer causal relationships at immune-cell resolution, revealing mechanisms not detectable in bulk analyses.

**Methods:** We conducted single-cell cis-eQTL MR across multiple immune cell subtypes to identify genes causally linked to lung cancer subtypes, including non-small cell lung cancer, further classified into adenocarcinoma and squamous cell carcinoma, and small cell lung cancer. Sensitivity and colocalization analyses confirmed shared genetic variants driving both expression and cancer risk. Bulk-tissue MR was conducted to see if the single-cell positive genes also show significance in whole blood and lung tissue. Phenome-wide MR (PheMR) further assessed pleiotropic associations across complex traits.

**Results:** Single-cell Mendelian Randomization (scMR) uncovered 375 cell-type specific causal links between gene expression and lung cancer subtypes. Colocalization confirmed shared causal variants in 35 gene-cell combinations. Cross-tissue validation highlighted five consistent genes—HSPA1B, HLA-DOA, GPX1, ZP3, and CORO1B. PheMR revealed these genes’ additional associations with other diseases, suggesting broader functional relevance.

**Conclusions:** The scMR workflow provides a high-resolution causal map of lung cancer susceptibility within immune contexts. The results nominate HSPA1B, HLA-DOA, GPX1, ZP3, and CORO1B as promising candidates for mechanistic and translational follow-up, demonstrating the power of single-cell causal inference in uncovering disease mechanisms obscured in bulk analyses.

## Introduction

Lung cancer remains a major cause of cancer morbidity and mortality worldwide (1). Although epidemiologic risk factors such as tobacco exposure and environmental pollutants are well established, germline variation also contributes meaningfully to inter-individual differences in susceptibility (2,3). Genome-wide association studies (GWAS) have identified numerous loci associated with lung cancer risk across histologic subtypes: non-small cell lung cancer (NSCLC) including adenocarcinoma and squamous cell carcinoma, and small cell lung cancer (SCLC) (4). However, most risk variants map to noncoding regions, and linkage disequilibrium (LD) often links many variants to each association signal (5,6). As a result, the path from a GWAS locus to the causal gene, relevant cellular context, and actionable biology remains uncertain for a considerable fraction of loci.

Transcriptomic quantitative trait loci (eQTLs) provide a principled route to connect noncoding variants to gene regulation. MR leverages eQTLs as genetic instruments to test whether genetically predicted changes in gene expression are causally related to disease risk (6). This approach may directly estimate the direction and magnitude of potential causal effects. Yet, many MR studies have been using bulk tissue eQTLs, which average signals across heterogeneous cell populations (7). When the effect of a variant is confined to a specific cell type or state, bulk averaging can attenuate or mask causal relationships. This is a critical limitation in the immune system, where cell-type specialization underpins function.

Advances in single-cell eQTL mapping now allow genetic effects on gene expression to be resolved within specific immune cell subsets, revealing that cellular context fundamentally shapes how genetic variation influences transcriptional regulation. Context-specific eQTLs thus emerge as key regulators of lung homeostasis and disease (8). This resolution is especially pertinent to lung cancer, where innate and adaptive immune cells influence carcinogenic progression through intricate molecular and signaling pathways (9,10). Accordingly, genetic regulation confined to specific immune subsets may shape lung cancer susceptibility in ways that become apparent only at single-cell resolution. Despite this rationale, the application of single-cell eQTL-based MR to lung cancer has been limited, and the extent to which cell-type specific regulation contributes causally to risk across histologic subtypes is unknown.

To address this gap, we hypothesized that germline variation influences lung cancer susceptibility through cell-type specific regulation of gene expression in immune cells, and that these effects can be uncovered by integrating single-cell eQTLs with lung cancer GWAS. We therefore performed a two-sample scMR analysis using immune-cell resolved eQTLs to estimate the causal impact of gene expression on risk across four lung cancer outcomes (overall NSCLC, adenocarcinoma, squamous cell carcinoma, and SCLC). Recognizing that MR alone cannot distinguish a shared causal variant from correlated signals in LD, we coupled scMR with pairwise conditional colocalization to test whether the same variant likely drives both the expression and disease association at each locus. Because disease-relevant regulation measured in single-cell may also manifest at the tissue level, we further evaluated convergent evidence using bulk eQTLs from whole blood and lung tissue. Finally, to situate lung cancer findings within a broader disease context and assess potential pleiotropy, we conducted a PheMR screen across hundreds of complex traits.

This multi-layered design of scMR discovery, conditional colocalization, cross-tissue MR, and PheMR provides a stringent causal filter that (i) limits false positives from LD, (ii) highlights the cellular compartments through which genetic risk is exerted, (iii) tests whether single-cell signals propagate to the tissue level, and (iv) differentiates disease-restricted from pleiotropic regulatory effects. Conceptually, the approach moves beyond the gene level to the resolution of gene-cell combinations to explore lung cancer mechanisms.

## Methods

### Study Design

The framework of this study is represented in Figure 1. Overall, the study aimed to use multiple public genomic databases to explore the causal relationship between gene and different types of lung cancer at single-cell level. We first extracted single cell eQTL data from OneK1K database for multiple types of immune cells and lung cancer GWAS data from FinnGen to conduct two-sample MR analysis. We further ran colocalization against significant gene-cell combinations from scMR result, narrowing down gene-cell combinations across different lung cancer subtypes. To further evaluate whether the genes from significance gene-cell combinations show positive results in lung and blood at the tissue level, we conducted a further two-sample MR using eQTLGen Consortium and GTEx database. Significant genes from scMR, colocalization, and bulk tissue MR were identified. To wrap up the analysis, we conducted a PheMR for the significant gene-cell combinations of the overlapping genes to identify their potential pleiotropic effects in other diseases. Figure 1 details this process.

**Figure 1.**
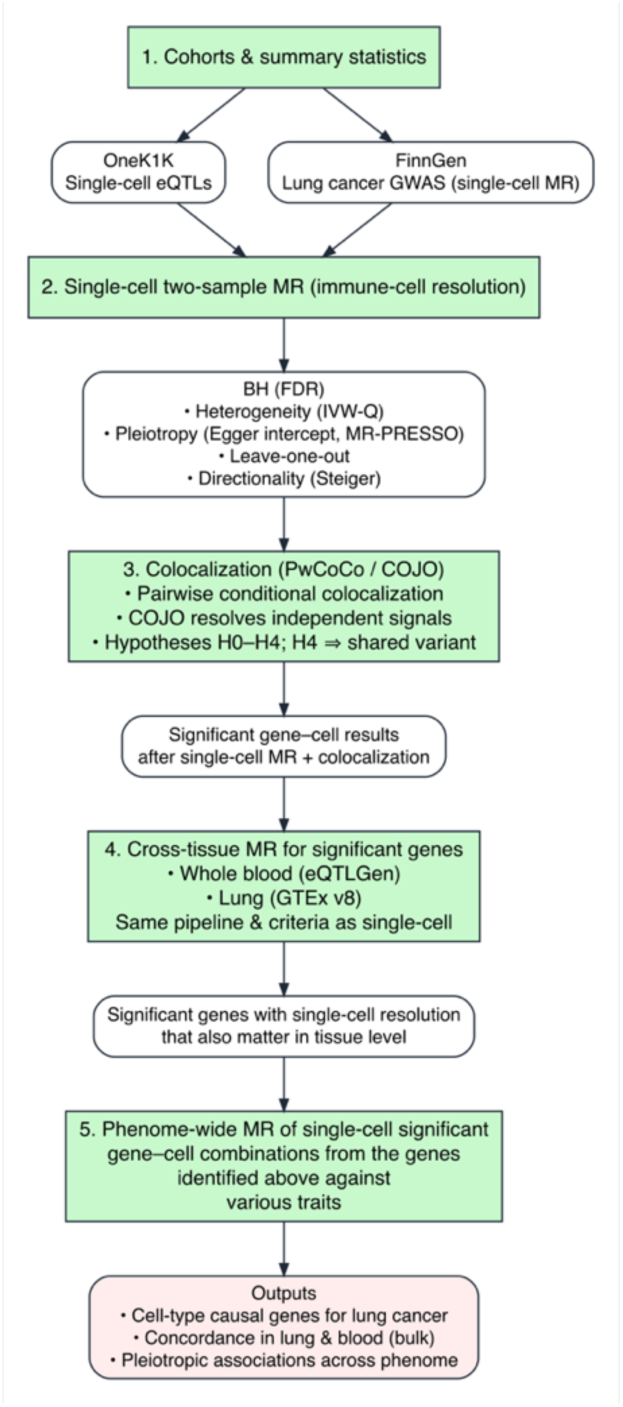
Flow diagram of study design.

### Data sources

Single-cell eQTL data were obtained from the OneK1K database, which profiled 1.27 million peripheral blood mononuclear cells from 928 individuals. The database identified 26,597 cis-eQTLs linking the expression of 16,597 genes across 14 immune cell types. The B cell lineage was subdivided into plasma cells (Plasma), immature/naïve B cells (B IN), and memory B cells (B Mem). CD4⁺ T cells were grouped into naïve and central memory (CD4 NC), effector and central memory (CD4 ET), and SOX4-expressing (CD4 SOX4) subsets. Similarly, CD8⁺ T cells were categorized into CD8 NC, CD8 ET, and S100B-expressing (CD8 S100B) populations. Innate immune cells were further distinguished into natural killer (NK) and NK-recruiting (NKR) cells, classical (Mono C) and non-classical (Mono NC) monocytes, and dendritic cells (DCs). (https://onek1k.org/) (11). Supplementary table 1 provides further specification. The eQTL summary statistics were provided by OneK1K and were used in this study as single-cell exposure data in the two-sample MR. These diverse immune cell types provided a comprehensive framework to explore the causal relationships between differential gene expression across immune subsets and lung cancer risk.

Bulk-level eQTL data were used to extend the single-cell analyses to broader tissue contexts. The whole blood eQTL data was obtained from the eQTLGen Consortium, which performed a large-scale meta-analysis of up to 31,684 blood samples collected across 37 cohorts to maximize statistical power for eQTL detection. This meta-analysis identified eQTL associations for 16,987 genes, providing a comprehensive map of genetic regulation in whole blood (https://www.eqtlgen.org/) (12). Lung tissue eQTL data were obtained from the Genotype-Tissue Expression (GTEx) v8 project, comprising 515 lung samples with matched genotype and RNA-seq profiles (https://www.gtexportal.org/home/downloads/adult-gtex/qtl) (13). Together, these datasets provided tissue-level gene expression for comparison and integrative analyses alongside the single-cell results. These data were used as bulk tissue exposure data in the two-sample MR.

GWAS summary statistics for lung cancer were obtained from FinnGen Release 12 (https://www.finngen.fi/en/access_results) (14). Four lung cancer subtypes were analyzed: NSCLC (N = 6446 cases, 378749 controls), adenocarcinoma (N = 2219 cases, 378749 controls), squamous cell carcinoma (N = 1898 cases, 379749 controls), and small-cell lung cancer (SCLC; N = 909 cases, 378749 controls). The curated GWAS datasets were used as outcome data for two-sample MR in single cell and bulk tissue analyses.

### Instrument selection

The MR framework uses genetic variants as instrumental variables (IVs) to infer the causal effect of an exposure, which is gene expression here, on an outcome, which is the lung cancer subtype. To yield valid causal estimates, the selected IVs must satisfy three core assumptions: [1] IVs are robustly associated with the exposure, [2] IVs are independent of any confounding factors, and [3] IVs influence the outcome solely through the exposure of interest (15).

Genetic instruments were derived from the OneK1K single-cell eQTL datasets and bulk tissue eQTL datasets (eQTLGen and GTEx v8). For each dataset, we extracted variants significantly associated with gene expression at a threshold of P < 1 × 10⁻⁵, focusing on cis-acting variants located within 1 Mb of the transcription start site to minimize horizontal pleiotropy (16). When multiple significant variants were available per gene-cell pair, we applied linkage disequilibrium (LD) clumping using PLINK to ensure independence, using an LD threshold of r² < 0.001 and a window size of 10 Mb based on the 1000 Genomes Project (European ancestry) reference panel (17,18). Instrument strength was assessed using the F-statistic, and weak instruments (F < 10) were excluded to prevent bias from underpowered variants. The remaining instruments were used to perform two-sample MR with harmonized GWAS data. Much of the analyses were conducted using the TwoSampleMR and ieugwasr R packages (19,20).

### Single-cell Mendelian Randomization and Sensitivity Analysis

We next performed two-sample MR (TwoSampleMR R package) to quantify the causal relationship between the gene expression of immune cells (exposure) and lung cancer subtypes (outcome) in each cell type (20). Effect estimates for eQTL instruments were harmonized with summary statistics from FinnGen Release 12, covering NSCLC in general, adenocarcinoma, squamous cell carcinoma, and SCLC. For each gene-cell-disease combination, the inverse-variance weighted (IVW) method under a random-effects model was applied when multiple independent variants were available, while the Wald ratio method was used for single-variant instruments. Significant causal effects were defined at a false discovery rate (FDR) < 0.05 by using the Benjamini-Hochberg method to adjust for p-values. The total number of tests and significant associations were summarized across 14 immune cell types and four lung cancer subtypes. Odds ratios (ORs) with 95% confidence intervals (CIs) were reported for all MR estimates.

We further assessed the robustness and validity of our Mendelian randomization results through a series of complementary sensitivity analyses. These included tests for consistency, heterogeneity, pleiotropy, and causal direction. Alternative estimators such as MR-Egger, weighted median, and weighted mode were applied to evaluate the consistency and stability of causal estimates under different model assumptions. Heterogeneity across instrumental variables was quantified using the IVW Q statistic, and potential horizontal pleiotropy was examined through the MR-Egger intercept and MR-PRESSO global test, with significant outlier variants excluded from downstream analyses (21,22). A nominal threshold of FDR < 0.05 indicated significance for heterogeneity and pleiotropy. Leave-one-out analyses were also performed to ensure that the overall effect was not driven by any single variant. Finally, causal directionality was verified using Steiger filtering, which compares the variance explained in exposure and outcome to detect potential reverse causation (23).

### Colocalization Analysis

To assess whether observed MR associations were driven by shared genetic architecture rather than LD between distinct causal variants, we performed pairwise conditional colocalization analysis (PWCoCo) (24). Colocalization evaluates five mutually exclusive hypotheses: H₀, no causal variants are present for either trait; H₁, a causal variant is associated with gene expression only; H₂, a causal variant is associated with disease only; H₃, both traits have causal variants within the same region but they are distinct; and H₄, both traits share the same causal variant. Evidence supporting H₄ indicates that the genetic signals for gene expression and disease risk are likely to arise from a shared causal variant rather than LD (25).

Unlike Bayesian colocalization methods such as coloc, PWCoCo performs conditional and joint (COJO) multi-SNP analysis to detect independent association signals within a genomic region. Each SNP is conditioned on the lead variant to identify conditionally independent signals, after which colocalization is tested for each pair of independent signals. This framework retains the single-variant assumption per test while accommodating multiple causal variants within the same locus (24). Colocalization was conducted for all SNPs within ±500 kb of each gene using prior probabilities (p₁ = p₂ = 1×10⁻⁵, p₁₂ = 1×10⁻⁷). Posterior probabilities were interpreted as the strength of evidence for each hypothesis, and a threshold of posterior probability of hypothesis 4 (PPH4) > 0.7 was considered strong evidence for colocalization between eQTL and GWAS signals.

### Bulk Tissue MR

To examine whether the causal effects identified in immune cell-specific MR analyses were also detectable at the bulk-tissue level, we conducted complementary MR analyses using eQTL summary statistics from the eQTLGen Consortium (whole blood) and GTEx v8 (lung tissue). The same analytical pipeline and significance criteria were applied as in the scMR analysis using TwoSampleMR. We specifically assessed whether genes showing significant causal effects in single-cell contexts also exhibited concordant direction and significance (FDR < 0.05) in blood and lung tissues. This cross-tissue MR analysis provided additional insight into how cell-type-specific regulatory mechanisms manifest at the tissue level, offering a broader perspective on transcriptional regulation underlying lung cancer susceptibility. The same suite of sensitivity analyses was performed as described for the scMR.

### PheMR Analysis

The purpose of our PheMR analysis was to systematically investigate causal associations between the identified genes and a broad spectrum of complex diseases, providing insight into their potential biological relevance beyond lung cancer. To accomplish this, we utilized genome-wide association summary statistics generated from SAIGE (Scalable and Accurate Implementation of Generalized Mixed Models), which analyzed more than 1,400 binary phenotypes from 408,961 European-ancestry participants in the UK Biobank (26) (https://www.leelabsg.org/resource). We selected 783 diseases with sample sizes greater than 500 from the SAIGE GWAS resource to maintain robust statistical power and minimize bias in causal inference. For each cell-gene-phenotype pair, Wald ratio MR analyses were performed using the corresponding lead cis-eQTL variant as the instrumental variable. Associations reaching FDR-corrected p < 0.05 were considered statistically significant.

## Result

### Single-cell Mendelian Randomization

After applying linkage disequilibrium (LD) clumping (r² < 0.001, 10 Mb window) and excluding weak instruments (F < 10), we retained 76,316 independent SNPs instrumenting 5,105 unique eGenes across 14 immune cell types as valid exposures. Using these eGenes, we performed scMR to test the causal effects of cell-type specific gene expression on lung cancer risk across four subtypes: NSCLC as a whole, further subtyping adenocarcinoma and squamous cell carcinoma, and SCLC. In total, 53,316 gene-cell-disease combinations were analyzed. After p-value adjustment, 375 associations (FDR < 0.05) remained significant, representing 184 unique genes across 14 immune cell subtypes. Most significant associations were enriched in CD4 NC cells, which accounted for 14.67% of all discoveries, while memory B cells yielded fewer associations, which accounted for only 2.4% of all discoveries.

When stratified by cancer subtype, 158, 31, 163, and 23 significant gene-cell combinations were detected for NSCLC, adenocarcinoma, squamous carcinoma, and SCLC, respectively (Figure 2). Out of the 375 significant associations, 113 gene-cell combinations present in ≥2 lung cancer subtypes. In particular, 110 of them displayed consistent causal directions across different lung cancer subtypes, suggesting shared genetic mechanisms underlying lung cancer susceptibility.

**Figure 2.**
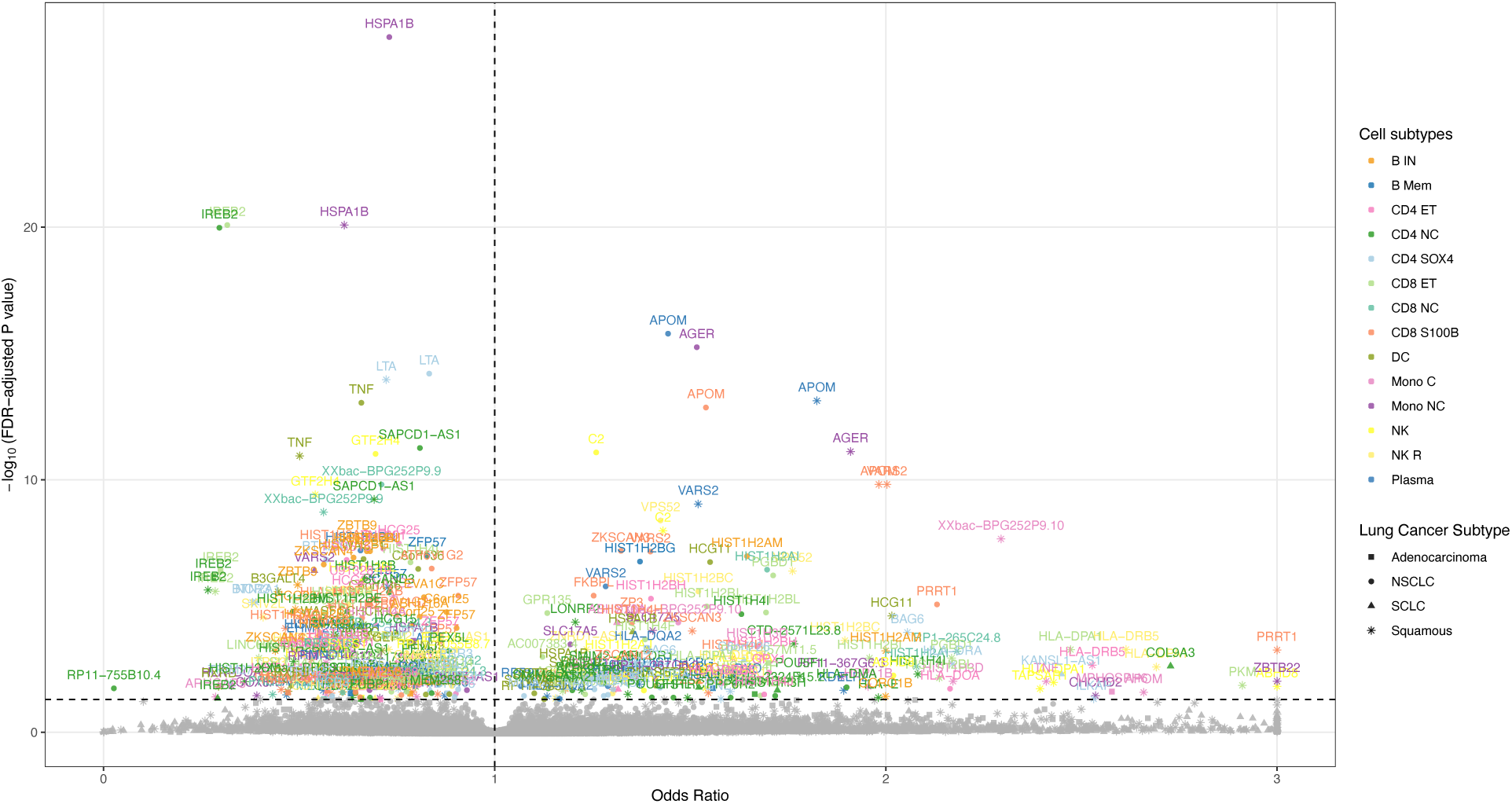
Volcano plot of the two-sample single-cell Mendelian Randomization results. Different colors correspond to different immune cell subtypes, and different shapes represent different lung cancer subtypes. Each point is labeled with its gene name. The abbreviations of the immune cell subtypes are defined in supplementary table 1.

### Colocalization and Bulk Tissue MR

To validate whether these scMR associations were driven by shared causal variants, we conducted PWCoCo colocalization for all significant gene-cell-disease pairs. From the 375 scMR significant gene-cell combinations across different lung cancer subtypes, we ran 2,909 colocalization tests as each combination’s eQTL signal may overlap with multiple loci of the lung cancer subtype. Of these, 35 pairs exhibited strong colocalization evidence (PPH4 > 0.7), involving 23 unique genes across 12 immune cell subtypes. When stratifying by lung cancer subtypes, there are 15, 6, 15, and 1 significant colocalization results in NSCLC, adenocarcinoma, squamous, and SCLC respectively (Figure 3).

**Figure 3.**
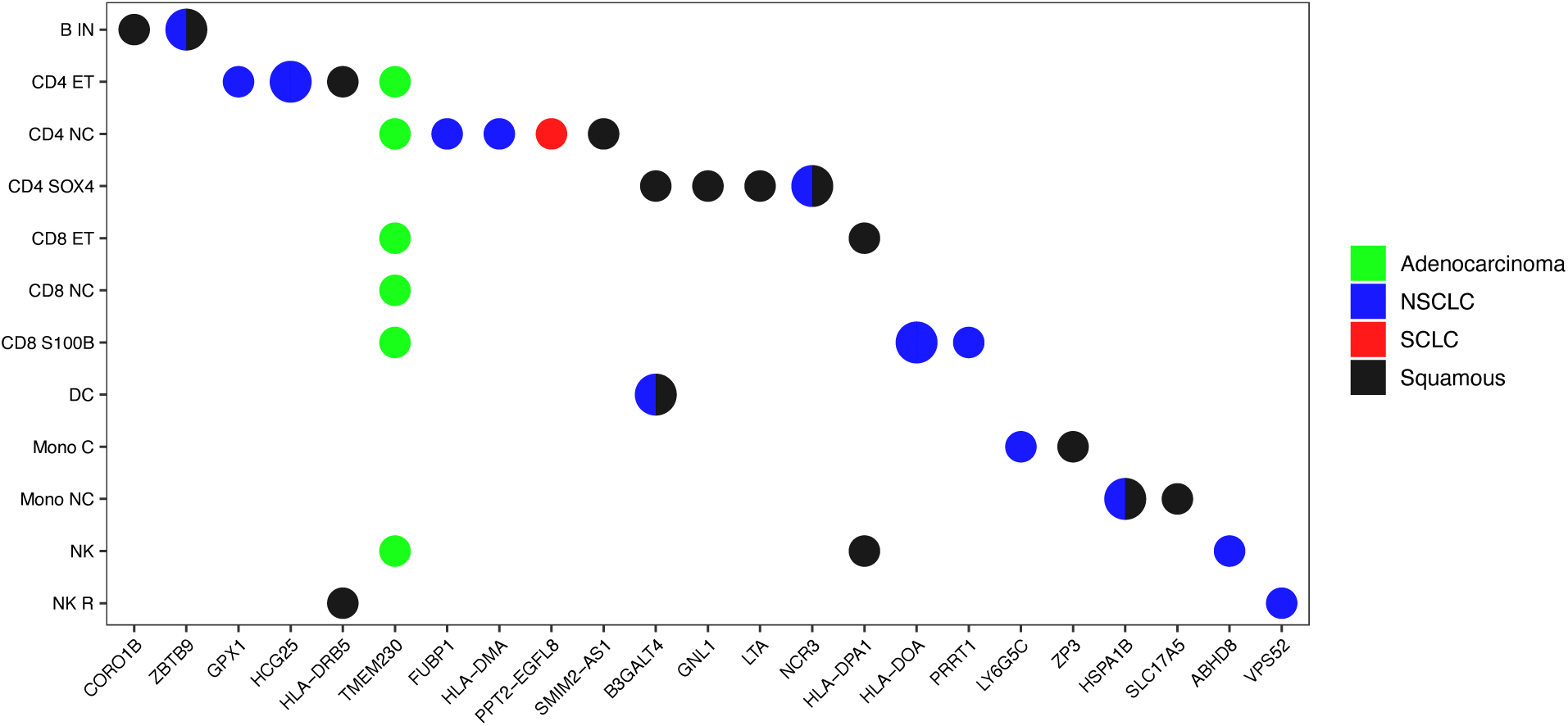
Heat map of the colocalization result. Genes are plotted in the x-axis and immune cell types are plotted in the y-axis. Color represents the lung cancer subtype. Each point represents a significant gene-cell combination’s presence in the corresponding lung cancer subtype. The size of each point is proportional to the number of significant colocalization results for that particular combination.

From the colocalization result, we then extended the analysis using bulk-tissue eQTL datasets to assess the significant gene-disease relationships in broader tissue contexts. The 23 unique genes filtered from colocalization is the candidate set. After applying identical instrument filtering, we retained 44 SNPs representing 19 unique genes from the eQTLGen dataset and 20 SNPs representing 13 unique genes from the GTEx v8 dataset. For blood-based MR using eQTLGen, 22 gene-disease pairs were tested, of which 3 remained significance (p-value < 0.05). For lung-based MR using GTEx v8, 16 gene-disease pairs were tested, of which 3 remained for the same criteria. Together, we found HSPA1B being significant in lung tissue for NSCLC (95% CI OR = 0.757–0.979, *p* = 0.0225) and squamous (95% CI OR = 0.597–0.954, *p* = 0.0187), HLA-DOA being significant in lung tissue in NSCLC (95% CI OR = 1.070–1.640, *p* = 0.0104), GPX1 being significant in blood in NSCLC (95% CI OR = 1.400–2.680, *p* = 0.000063), and ZP3 and CORO1B being significant in blood for squamous (95% CI OR = 0.475–0.799, *p* = 0.000253; 95% CI OR = 1.300–2.550, *p* = 0.000484) (Figure 4A).

**Figure 4A.**
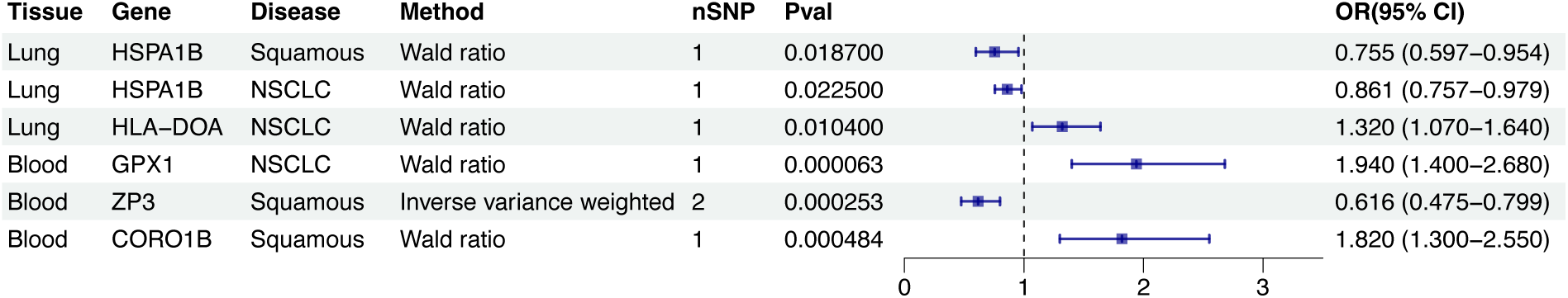
Forest plot showing bulk-tissue MR estimates for the causal effects of the selected genes on lung cancer subtypes using eQTL instruments from GTEx v8 lung and eQTLGen whole blood. Each interval represents an OR with a 95% confidence interval for one gene-subtype pair.

At this stage, we identified five genes whose causal effects were consistently supported across multiple analytic layers—scMR, colocalization, and at least one bulk-tissue MR—indicating that their associations were unlikely to arise from linkage or dataset-specific artifacts. These genes, HSPA1B, HLA-DOA, GPX1, ZP3, and CORO1B, demonstrated reproducible significance across both immune-cell and tissue-level contexts, providing high-confidence candidates for lung cancer susceptibility.

To further clarify how these cross-level signals arise, we traced each validated gene back to its originating single-cell associations found from scMR and proved by colocalization, thereby identifying the specific immune cell types in which gene expression most strongly influenced cancer risk. This mapping revealed six significant distinct gene-cell-disease combinations (Figure 4B). Increased HSPA1B expression in non-classical monocytes was protective in both NSCLC (95% CI OR = 0.693–0.771, FDR = 2.95×10⁻²⁸) and squamous cell carcinoma (95% CI OR = 0.559–0.677, FDR = 8.24×10⁻²¹), indicating a robust, cross-histology effect. In contrast, GPX1 expression in CD4⁺ effector T cells increased NSCLC risk (95% CI OR = 1.322–2.189, FDR = 0.004308), while HLA-DOA expression in S100B⁺ CD8⁺ T cells showed a protective effect (FDR = 0.017193, 95% CI OR = 0.621–0.874). Similarly, higher ZP3 expression in classical monocytes was protective in squamous carcinoma (95% CI OR = 0.414–0.783, FDR = 0.032696), whereas increased CORO1B expression in immature/naïve B cells elevated squamous risk (FDR = 0.037846, 95% CI OR = 1.356–2.947).

**Figure 4B.**
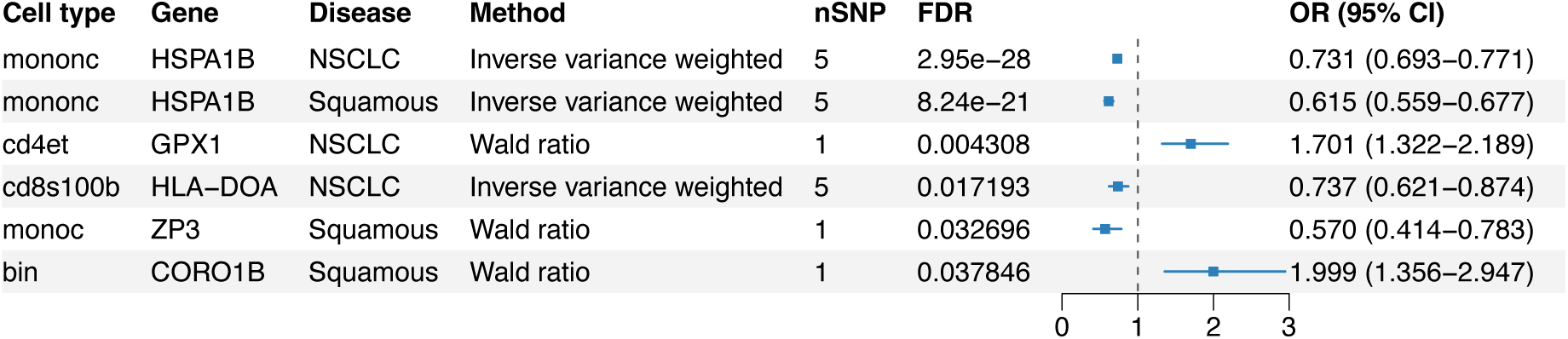
Forest plot summarizing the selected genes’ scMR results of the significant gene-cell-disease combinations that were validated by colocalization (PPH4 > 0.7). Each interval represents the causal effect estimate (OR, 95% CI) of gene expression in a specific immune cell type on lung cancer risk, illustrating the cell-type resolved genetic architecture of susceptibility.

Taken together, these findings delineate a continuum from cell-type-specific regulation to tissue-level manifestation, highlighting how germline variants exert disease effects through distinct immune compartments. The genes collectively span diverse biological functions: HSPA1B in protein stress response, HLA-DOA in antigen presentation regulation, GPX1 in oxidative balance, ZP3 as a regulatory proxy, and CORO1B in cytoskeletal remodeling, illustrating multiple mechanistic routes through which immune regulation contributes to lung cancer susceptibility (27–32).

### Phenome-Wide Mendelian Randomization Analysis

Building on the validated single-cell and bulk-tissue results, we next examined whether these five causal genes, HSPA1B, HLA-DOA, GPX1, ZP3, and CORO1B, also influence other complex traits beyond lung cancer. We performed a PheMR analysis across 783 diseases grouped into 17 major categories, using genome-wide association summary statistics from the UK Biobank SAIGE resource. For each overlapping gene, we selected its corresponding significant cell types from the scMR layer, generating an initial set of 33 SNPs representing 11 unique gene-cell combinations. After filtering for valid, independent instruments available in the PheMR database, seven gene-cell pairs were retained as exposures for downstream analysis: GPX1 in CD4 ET, CORO1B in B IN, HSPA1B in Mono C, ZP3 in Mono C, HSPA1B in Mono NC, HSPA1B in DC, and ZP3 in CD8 S100B. HLA-DOA was excluded because its extensive linkage disequilibrium within the HLA locus prevented reliable instrument definition (33). Associations reaching FDR < 0.05 were considered statistically significant (Figure 5).

**Figure 5.**
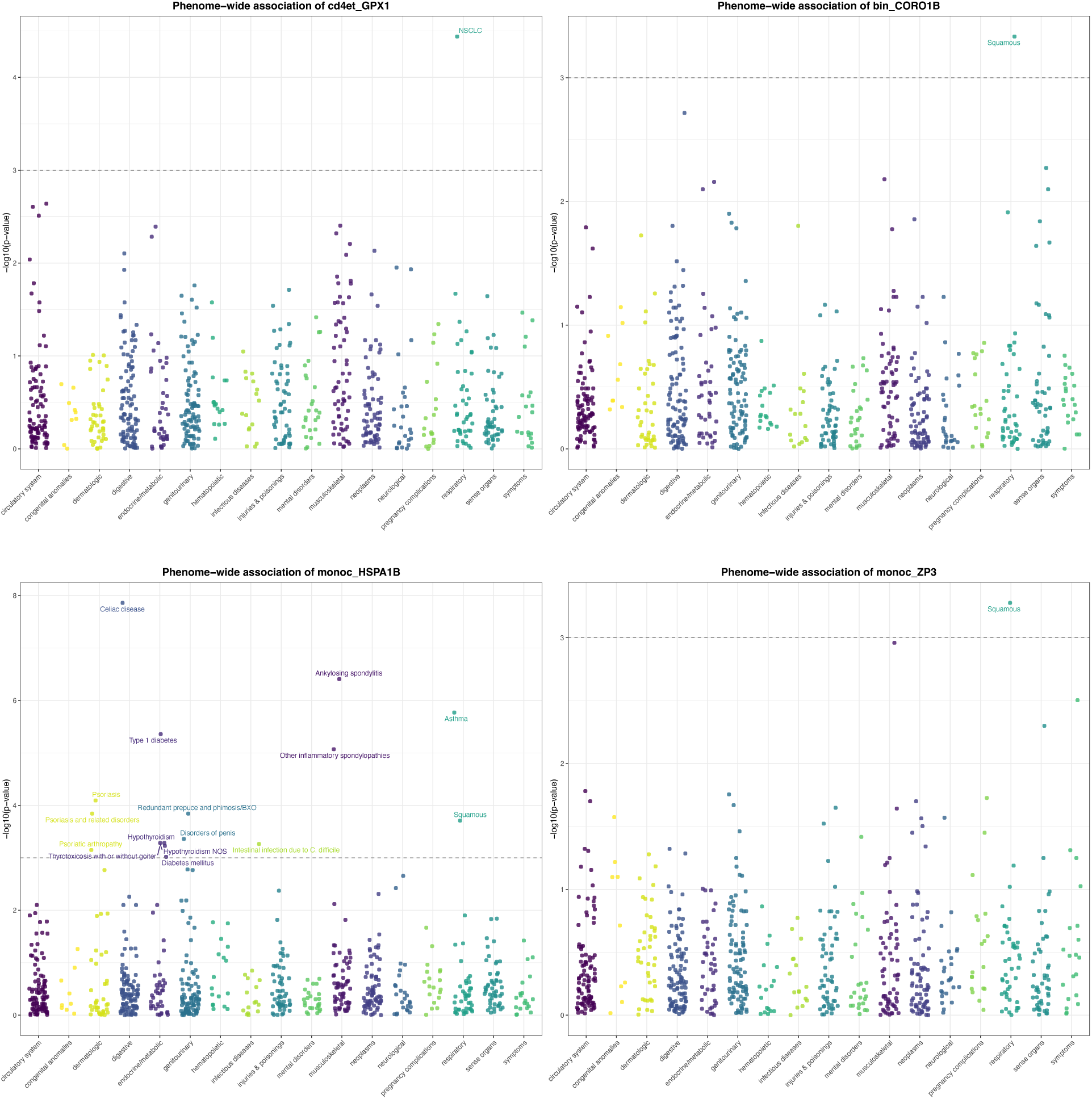
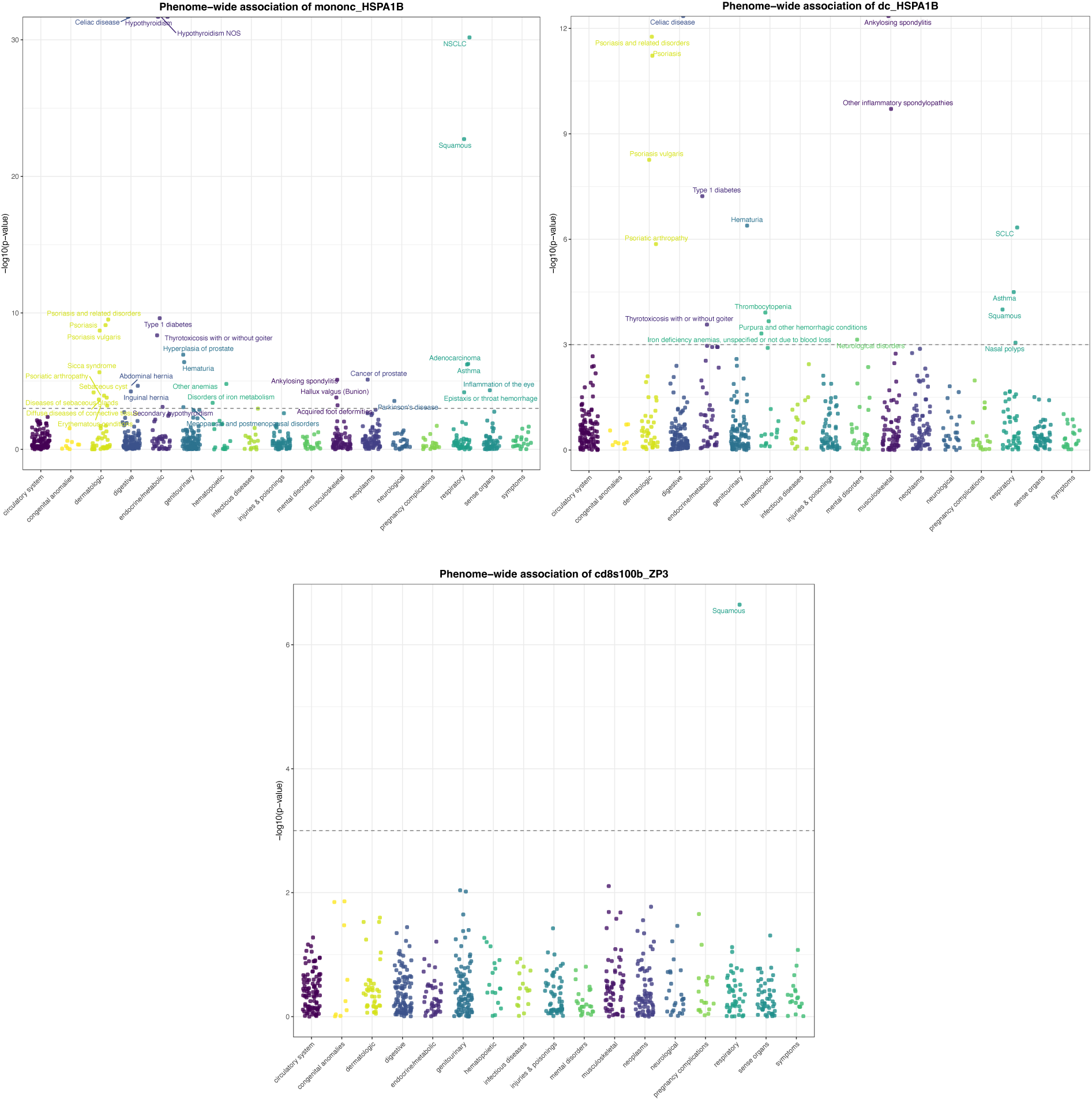
PheMR result of the selected gene-cell combination on different diseases. Each bar represents a broad disease category, and each point represents a specific disease within the category.

This phenome-wide screen extended our analysis beyond lung cancer, assessing whether the same genetically predicted expression changes that modulate cancer risk also contribute to other disease processes. The results revealed that HSPA1B was the only gene showing additional significant causal associations outside of lung cancer, and these occurred consistently across all three of its tested corresponding cell types, Mono C, Mono NC, and DC. In contrast, other gene-cell combinations exhibited significance only for their original lung cancer subtypes from the scMR results. Together, these findings suggest that HSPA1B acts as a broadly pleiotropic stress-response gene influencing multiple physiological and pathological states, whereas GPX1, ZP3, and CORO1B exert more context-restricted, lung-specific effects.

## Discussion

To our knowledge, this study is the first to apply Mendelian Randomization at single-cell resolution to reveal causal relationships between gene expression and lung cancer subtypes. By integrating scMR, colocalization, bulk-tissue MR, and PheMR, we delineate immune cell-resolved causal mechanisms for lung cancer risk that are largely obscured in bulk analyses. From 53,316 gene-cell-subtype tests, we identified 375 FDR-significant associations, of which 35 gene-cell-disease pairs showed strong colocalization (PPH4 > 0.7). Cross-tissue validation in whole blood and lung tissue further converged on five genes—HSPA1B, HLA-DOA, GPX1, ZP3, and CORO1B—as robust, immune-anchored candidates that exert wider influences at the tissue level. At the single-cell level, these signals localize to distinct immune compartments, including non-classical and classical monocytes, CD4⁺ effector T cells, S100B⁺ CD8⁺ T cells, and immature or naïve B cells, underscoring how germline regulation of specific leukocyte programs contributes to lung cancer susceptibility.

A key conceptual advance of this study is that single-cell MR uncovers causal relationships that bulk tissue analyses systematically miss. We identified 23 unique genes among the 35 gene-cell-disease pairs supported by both scMR and colocalization. However, 18 genes were missing or negative in bulk-tissue MR, indicating that these genetic effects manifest only within specific immune environments rather than at the tissue-wide average. This highlights that many regulatory mechanisms are fundamentally cell-state dependent, becoming detectable only when the relevant cellular context is isolated. For the remaining five genes that did replicate in bulk tissue, scMR adds an essential layer of mechanistic clarity revealing the exact immune subsets in which these causal signals arise, resolving biological questions that bulk-tissue MR cannot answer. This is important because although germline causal SNP exists in every tissue, only certain cell types have the transcriptional machinery to convert that variant into functionally meaningful consequences (34,35). Thus, scMR not only expands discovery but also localizes the source of bulk associations, yielding a more coherent, cell-centric understanding of lung cancer risk. From the 35 significant associations, we then focused on the subset of genes that were also validated in bulk-tissue MR, where single-cell resolution allows us to pinpoint the precise immune compartments through which these broader tissue-level effects arise.

From our single-cell results, protective HSPA1B expression in non-classical monocytes likely reflects adaptation to the hypoxic, nutrient-poor, and oxidative stresses characteristic of the NSCLC microenvironment. Such conditions strongly induce HSPA1B, enabling monocytes to maintain proteostasis and sustain inflammatory activity under stress (36,37). HSPA1B also encodes for heat-shock protein 70 (Hsp70), which is frequently displayed on the membrane of NSCLC cells and released in lipid vesicles, with particularly high levels in squamous tumors. Although membrane Hsp70 correlates with tumor aggressiveness, it simultaneously serves as a potent immunogenic ligand: NK cells pre-stimulated with Hsp70-derived TKD peptide plus IL-2 upregulate cytotoxic receptors and selectively kill mHsp70⁺ tumor cells. (38). Besides, extracellular Hsp70 activates monocytes directly, inducing IL-1β, IL-6, TNF-α and complement pathways, thereby amplifying innate immune responses (39). More broadly, Hsp-family proteins facilitate cancer immunosurveillance by presenting chaperoned tumor peptides to monocytes and dendritic cells via CD40, CD91, and LOX-1, enhancing antigen cross-presentation (40). Taken together, these findings support a model in which genetically increased HSPA1B expression strengthens monocyte stress tolerance and antigen-handling capacity in NSCLC, particularly in squamous carcinoma, contributing to a more immunogenic microenvironment and reduced disease risk.

In contrast, GPX1, which encodes the ubiquitously expressed glutathione peroxidase-1, is a key intracellular antioxidant enzyme that reduces hydrogen peroxide and lipid hydroperoxides using glutathione. In T cells, a transient rise in reactive oxygen species (ROS) is required for early T-cell receptor signaling and clonal expansion, whereas GPX1 acts to quench these activation-induced ROS bursts. Experimental models show that GPX1-deficient T cells exhibit elevated ROS, increased IL-2 production, and accelerated proliferation, indicating that excessive GPX1 activity can suppress ROS-dependent activation pathways. Beyond the immune context, GPX1 overexpression is common across many cancers and contributes to chemoresistance and metabolic stress tolerance, including protection from cisplatin-induced cytotoxicity and support of oxidative phosphorylation under nutrient deprivation (41–43). Clinical data further suggest that high GPX1 expression or GPX1 genetic variants can associate with poorer outcomes or increased susceptibility in several malignancies (44,45). Taken together, these findings support a model in which elevated GPX1 in CD4⁺ effector T cells lowers activation-associated ROS signaling, restrains effector function, and ultimately contributes to a more immunosuppressive environment, consistent with the risk-increasing effect we observe for GPX1 in CD4⁺ effector T cells in NSCLC.

Our finding that HLA-DOA expression in CD8⁺ S100B⁺ T cells associates with a protective effect in lung cancer suggests a convergence between non-classical MHC-II regulation and a stress-adapted cytotoxic T-cell phenotype. CD8⁺ S100B⁺ lymphocytes represent a distinct effector subset marked by inducible S100B expression, which rises under hypoxic conditions and mediates immune–microenvironmental crosstalk (46,47). Although S100B has been linked to hypoxia-mediated activation, EMT pathways, and immune infiltration in tumors, circulating S100B⁺ lymphocytes are predominantly CD3⁺CD8⁺ T cells that release S100B upon stimulation, potentially activating local myeloid cells (46). The presence of HLA-DOA, a non-classical MHC-II regulator that counterbalances HLA-DM to enable presentation of DM-sensitive antigens, may endow these CD8 T cells with atypical antigen-processing functions or reflect an APC-like transcriptional program frequently observed in tumor-reactive CD8 populations (28). Because tumors frequently escape immunity through downregulating classical HLA expression, higher HLA-DOA in these CD8⁺ S100B⁺ cells could enhance detection of otherwise poorly presented peptides, supporting sustained anti-tumor immunity in immune-suppressive niches such as hypoxic lung tumors (48).

The detection of protective ZP3 expression in classical monocytes is striking given that ZP3 is classically an oocyte-restricted glycoprotein essential for zona pellucida formation and sperm-egg recognition (49). However, recent studies revealed that ZP3 is aberrantly expressed in multiple malignancies, including lung cancer, where it generates intracellular isoforms that may serve as tumor-associated neoantigens (31). Rather than indicating a pro-tumoral program, ZP3 expression in monocytes may reflect heightened antigen uptake and immune surveillance toward ZP3-expressing tumor cells. Monocytes readily internalize tumor-derived proteins and RNA through phagocytosis, exosomes, and membrane exchange. Intracellular ZP3 accumulation could therefore mark active monocyte engagement with malignant cells, leading to enhanced antigen processing and presentation.

Lastly, our analysis identifying CORO1B expression in naïve B cells as causally increasing squamous risk adds a new immune-cell context to a protein previously characterized almost exclusively through its cytoskeletal functions. CORO1B is a conserved actin-binding protein that regulates Arp2/3-branched actin networks, lamellipodial protrusion dynamics, and actin turnover (32,50). In multiple cellular systems, loss of CORO1B leads to increased branched actin density, reduced actin turnover, and altered protrusive behavior, establishing it as a key regulator of actin-dependent structural organization (50). Beyond its role in protrusion dynamics, CORO1B also participates in junctional architecture, colocalizing with VE-cadherin and interacting with integrin-linked kinase and α-parvin in endothelial cells (32). Although prior literature has not examined Coro1B specifically in naïve B cells or in cancer immunity, CORO1B does appear in B-cell specific epigenetic analyses, where it is hypermethylated in B cells relative to T cells and monocytes, demonstrating potential lineage-restricted regulation (51). Within this framework, our causal inference result indicates that CORO1B expression in naïve B cells has measurable relevance to cancer, extending the functional reach of a well-established cytoskeletal regulator into a new immunological and oncologic context.

At a broader level, several conceptual themes emerge. First, cellular resolution matters. Many causal effects were confined to specific immune subsets and were diluted or undetectable in pure bulk analyses, demonstrating that disease-relevant regulation often operates within narrowly defined cellular contexts. Also, conditional colocalization is essential to distinguish true causal overlap from linkage-driven correlations, as only a small proportion of MR-significant pairs achieved strong PPH4 support. The multi-layered framework adopted here, scMR followed by PWCoCo, cross-tissue MR, and PheMR, acts as a rigorous causal filter that progressively narrows candidates to those supported by convergent genetic evidence. The PheMR analysis further distinguished pleiotropic from disease-restricted effects. HSPA1B displayed additional causal links to non-cancer traits, consistent with its broad role in proteostasis, whereas HLA-DOA, GPX1, ZP3, and CORO1B appeared more lung-specific. This distinction offers a useful prioritization framework in translational contexts, where broadly pleiotropic genes may represent fundamental cellular regulators but carry greater off-target risk, while context-specific genes provide clearer hypotheses for mechanism-of-risk and targeted intervention.

Several limitations warrant consideration. Differences in ancestry and linkage disequilibrium between the OneK1K, FinnGen, and reference datasets may influence instrument selection and colocalization resolution, particularly in complex regions such as HLA. The sample size of the single-cell eQTL resource, though large for this design, may still limit the power for lowly expressed genes and rare immune states. Because the single-cell eQTLs were collected from peripheral blood mononuclear cells, they primarily capture systemic immune regulation rather than lung-resident or tumor-infiltrating immune programs and thus may miss context-specific effects occurring within the pulmonary microenvironment (11). Residual horizontal pleiotropy cannot be fully excluded despite sensitivity analyses, and single-variant Wald ratio estimates used for some genes remain sensitive to instrument validity. Gene attribution in loci such as ZP3 and HLA-DOA also requires experimental confirmation. Finally, PheMR relied on available UK Biobank outcomes predominantly from individuals of European ancestry, and null findings may reflect limited power or incomplete phenotype coverage.

Future work should aim to replicate these associations in independent single-cell eQTL datasets, perform fine-mapping to resolve causal variants, and integrate single-cell chromatin QTL and perturb-seq data to connect variants with regulatory elements and downstream gene networks. Multivariable MR could disentangle co-regulated transcripts, and stratified or gene-environment MR analyses by smoking status, sex, or ancestry could illuminate context-dependent effects. Functionally, targeted perturbations, such as modulating ZP3 in classical monocytes and CORO1B in immature B cells, will be essential to confirm mechanistic causality and define how immune regulation translates into cancer risk from unexpected angles.

In conclusion, this study demonstrates that scMR combined with conditional colocalization and cross-tissue validation can reveal immune mechanisms of lung cancer susceptibility that remain hidden in bulk tissue analyses. The convergence on HSPA1B, HLA-DOA, GPX1, ZP3, and CORO1B underscores how germline variation in distinct immune lineages contributes to tumor initiation and progression through multiple biological pathways. More broadly, our results establish cell-type resolved causal inference as a powerful approach for linking genetic regulation to disease, providing a framework that can be generalized to other complex traits where cellular context defines genetic effect.

## Data Availability

All data produced in the present study are available upon reasonable request to the authors

https://onek1k.org/

https://www.eqtlgen.org/

https://www.gtexportal.org/home/downloads/adult-gtex/qtl

https://www.finngen.fi/en/access_results

https://www.leelabsg.org/resource

## Acknowledgements

This work was supported by the Donald and Elizabeth Cooke Distinguished Professorship of Cancer Research held by Dr. Xiao-Fan Wang at the Duke University School of Medicine. Qimao Yang received additional support from the 2025 Biology Summer Internship/Experience Program (BioSIP) administered by the Cornell University OSice of Undergraduate Biology.

