## Supplemental Table 1 for "Single-Cell Mendelian Randomization Identifies Cell-Type Specific Genetic Drivers of Lung Cancer Subtypes"

### Supplementary Material

Supplementary Table 1. Cell type abbreviation and definition.

| Cell type abbreviation | Cell type | Summarized classification |
|---|---|---|
| CD4 ET | CD4+ KLRB1+ T cell | $CD4^+$ effector/central memory T cell |
| CD4 NC | CD4+ KLRB1- T cell | $CD4^+$ naïve/central memory T cell |
| CD4 SOX4 | CD4+ SOX4+ T cell | $CD4^+$ SOX4-expressing T cell |
| CD8 NC | CD8+ LTB+ T cell | $CD8^+$ naïve/central memory T cell |
| CD8 ET | CD8+ GNLY+ NKG7+ T cell | $CD8^+$ effector/central memory T cell |
| CD8 S100B | CD8+ S100B+ T cell | $CD8^+$ S100B-expressing T cell |
| NK | XCL1- NK | NK cell |
| NK R | XCL1+ NK | NK-recruiting cell |
| B Mem | TCL1A- FCER2- B cell | Memory B cell |
| B IN | TCL1A+ FCER2+ B cell | Immature/naïve B cell |
| Plasma | IgJ+ B cell | Plasma cell |
| Mono C | Monocyte CD14+ | Classical monocyte |
| Mono NC | Monocyte FCGR3A+ | Non-classical monocyte |
| DC | Dendritic cell | Dendritic cell |
